# Deep learning Model for Recognizing Monkey Pox based on Dense net-121 Algorithm

**DOI:** 10.1101/2022.12.20.22283747

**Authors:** Mohamed Torky, Ali Bakheit, Mohamed Bakry, Aboul Ella Hassanien

**Author notes:** Scientific Research Group in Egypt (SRGE), http://egyptscience.net/.

## Abstract

While the world is trying to get rid of the Covid 19 pandemic, the beginning of the monkeypox(MPX) pandemic has recently appeared and is threatening many countries of the world. MPX is a rare disease caused by infection with the MPX virus, and it is among the same family of pox viruses. The danger is that MPX causes pustules all over the body, which causes a revolting view to the body regions and works as a source of infection in case of skin contact between individuals. Pustules and rashes are common symptoms of many pox viruses and other skin diseases such as Measles, chicken pox, syphilis, Eczema, etc, Therefore, the medical and clinical diagnosis of monkeypox is one of the great challenges for doctors and specialists. In response to this need, Artificial intelligence can develop aid systems based on machine and deep learning algorithms for diagnosing these types of diseases based on datasets of skin images to those types of diseases. In this paper, a deep learning approach called Dense Net-121model is applied, tested, and compared with the convolution neural network (CNN) model for diagnosing monkeypox through a skin image dataset of MPX and Measles images. The most significant finding to emerge from this study is the superiority of the Dense Net-121 model over CNN in diagnosing MPX cases with a testing accuracy of 93%. These findings suggest a role for using more deep learning algorithms for accurately diagnosing MPX cases with bigger datasets of similar pustules and rashes diseases.

## 1. Introduction

While many countries of the world are still trying to eradicate the COVID-19 pandemic and its huge economic losses, the monkeypox (MPX) pandemic has emerged in the face of the world as another contagious viral disease no less dangerous than COVID-19 and other epidemic diseases. MPX is a viral infection disease where its symptoms appear after a week or two with fever and other non-specific symptoms, then rash with lesions are produced at various body regions that usually last for 2–4 weeks before drying up.

The MPX was confirmed as an ongoing pandemic in May 2022. The first case was detected on 6 May 2022 in a passenger on a travel flight to Nigeria. On 23 July, the Director-General of the World Health Organization (WHO), announced the MPX outbreak as a public health emergency of international concern **[1]**. The pandemic specified the first time MPX has spread widely in the west and the center of Africa, but from 18 May onward, Europe and America becomes increasingly the regions of endemic to MPX. Since the beginning of the MPX outbreak and as of 29 August 2022, several confirmed MPX cases passed 18 072 cases from 29 EU/EEA countries **[2]**. In the USA, it is reported that the number of confirmed MPX cases passed 10,768, according to the center for disease control and prevention (CDC) data and the number is increasing **[3]**. Globally, the number of confirmed MPX cases reached 50,531 cases in over 100 countries, and the USA become the highest number of MPX cases in the world **[4]**. Since the infection rate is based on skin contact between individuals or animals, the MPX spread rate is low compared to COVID-19 and other infectious diseases. However, its danger is that it causes pustules all over the body, which cause a revolting view to the body regions and work as a source of infection. Some reports and studies. Many studies and reports have suggested that the cause of MPX infection is due to sexual intercourse between homosexuals **[5**,**6]**. Therefore, scientists and all public health authorities have to update their MPX communication strategies to more strongly undermine homosexual activity and limit their whereabouts.

Skin bumps and rashes are a common feature of many diseases besides MPX, for instance, chicken pox, syphilis, and Measles. Therefore, clinical diagnosis of MPX is in an early stage and challenging due to its similarity with rash and skin lesion-based disease. Hence, recognizing MPX cases through Skin bumps and rashes images using Artificial Intelligence (AI) is an important research problem **[7]**. In situations, where confirmatory Polymerase Chain Reaction (PCR) tests are not available easily, AI-based detection of MPX skin lesions could be effective for surveillance and fast recognition of suspected cases. Deep learning methods have been found effective in the automated detection of skin lesions, providing the availability of sufficient training instances. It is well known that AI approaches are data-driven, which require a huge number of data samples for training and developing AI models effectively. However, there are no publicly available and dependable datasets of MPX in the form of a digital images of rashes or skin lesions. The researchers tackled this issue by employing a web-scrapping tool for collecting digital skin lesion images of MPX and similar rashes-based diseases to develop efficient AI-based MPX recognition models **[8]**. To our knowledge, few research efforts have been made for designing AI-based MPX identification algorithms and most of this work has been published as preprints **[9**,**10]**. However, these algorithms did not achieve accuracy better than 82% in detecting MPX cases.

In response to this shortcoming, this study tries to find an alternative solution to recognizing the MPX problem with better accuracy. In this paper, two algorithms, Dense net-121, and Convolution Neural Network (CNN) have been applied and compared on a developed dataset including MPX and Measles images **[9]**. The implementation results proved the superiority of the Dens net 121 algorithm compared to the CNN model in detecting MPX images.

The remaining sections of this paper can be organized as follows: Section 2 presents the literature review, section 3 discusses the proposed method, section 4 presents and discuss the implementation results, and finally, section 5 concludes and summarizes this work.

## 2. Literature Review

The literature on detecting MPX disease has highlighted some tries of the machine and deep learning models based on skin image datasets.

Krishna et al **[11]** developed a hybrid deep learning model based on CNN and Long short-term memory (LSTM) algorithms for detecting sentiment polarities on Monkeypox tweets through Twitter s. The implementation results clarified the prediction accuracy of 94% detecting sentiment polarities on Monkeypox using the monkeypox tweet dataset. These results have been compared with other machine learning models and showed superiority to the proposed deep learning model.

Chiranjibi Sitaula and Tej B Shahi **[12]** compared 13 pre-trained deep learning models for detecting the MPX virus. The authors validated these models based on a public dataset. The validation results showed that the used deep learning algorithms achieved an average MPX detection accuracy of 87.13%. The authors claim that these findings are encouraging to be applicable and assist health practitioners.

Some studies used fuzzy logic with neural networks to detect the MPX virus. Tom et al **[13]** proposed a Neuro-Fuzzy Model for recognizing MPX disease. The proposed system can be used as a recommendation system for doctors and paramedical health workers for diagnosing MPX disease as it doesn’t require training while implementing its methodology. Moreover, young medical practitioners can use it as a decision support system to acquire more knowledge on the diagnosis of MPX.

Luis Muñoz-Saavedra et al **[14]** developed MPX diagnostic-aid system using convolutional neural networks (CNN) and a skin image dataset. The authors compared the performance of using the CNN model and CNN integrated with ResNet50, EfficientNet-B0, and MobileNet-V2. The obtained results showed that the last ensample models achieved an accuracy of 98% compared with using the CNN model alone, which achieved an accuracy of 93% in detecting MPX disease within a skin image dataset consisting of normal, MPX, and other skin diseases.

Although the provided works represent serious steps toward developing an AI-based system for diagnosing MPX diseases, more research efforts are still required for developing a more accurate AI system for recognizing MPX based on blood or skin datasets to help doctors to recognize MPX compared to other skin diseases such as Measles, Syphilis, chickenpox, etc.

## 3. MPX Recognition Model

AI approaches such as machine and deep learning can be used also for diagnosing many skin and pox family diseases by recognizing the key symptoms of each one. AI-based methods provide a means of classifying features of a pox disease, then recognizing major features, which can be used to diagnose pox disease types such as Measles, Syphilis, chickenpox, or monkeypox (MPX). A proposed machine learning-based model is developed to recognize MPX using a modified Dense net-121 model and skin images dataset including MPX and Measles images **[9]**. Table 1 describes the used dataset, and Figure 1 depicts samples of this dataset. Figure 2 depicts the architectural design model of the proposed deep learning approach based on the Dense net-121 model. The main motivation behind the use of DenseNet-121 is that it can improve feature reuse, mitigates the problem of vanishing gradient, and reduce parameter usage, which is useful for training deep learning models. Moreover, DenseNet-121 has proven effective in diagnosing diseases based on image datasets **[15]**.

**TABLE 1.** Dataset description

| Training Data |  | Testing Data |  |
| --- | --- | --- | --- |
| Number of Measles images | 19 | Number of Measles images | 8 |
| Number of MPX images | 35 | Number of MPX images | 8 |

**Fig.1.**
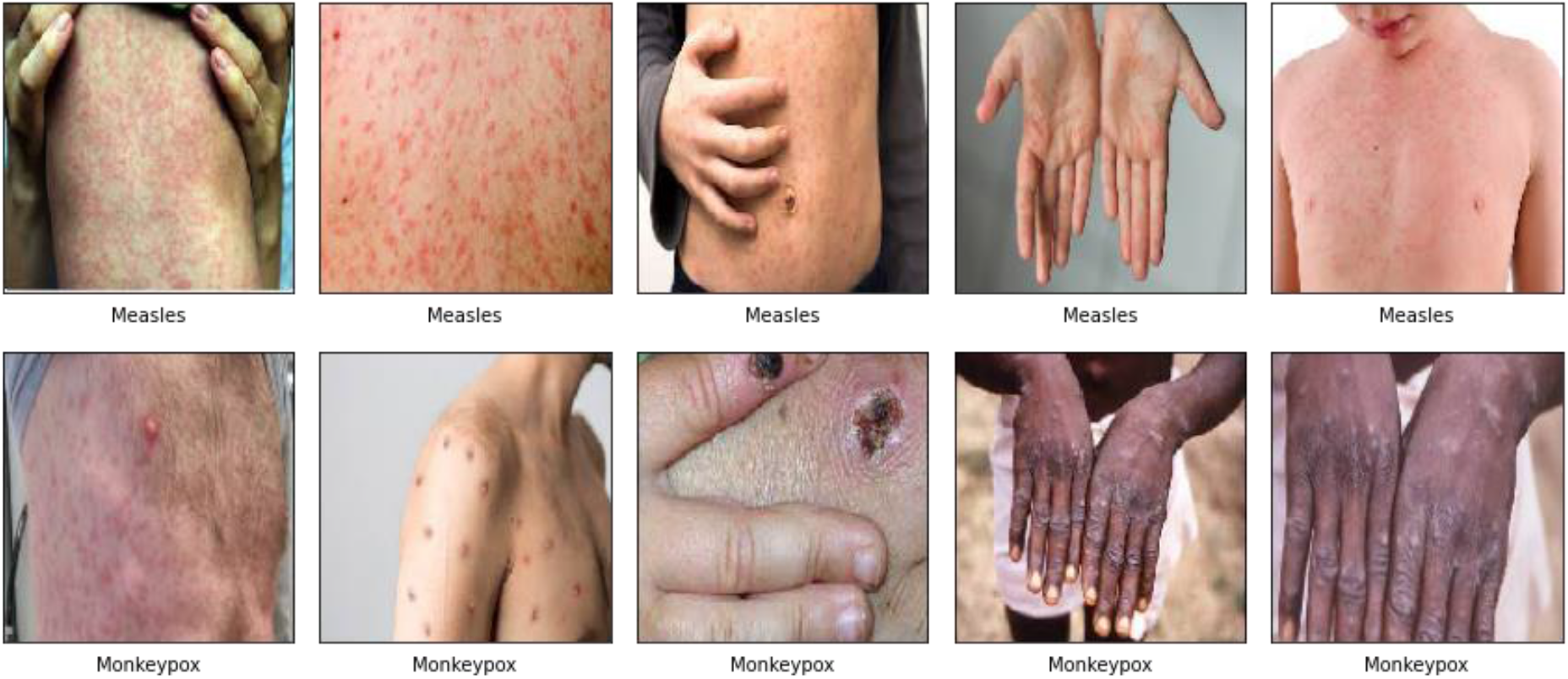
Samples of the used dataset: Measles, and MPX images

**Fig.2.**
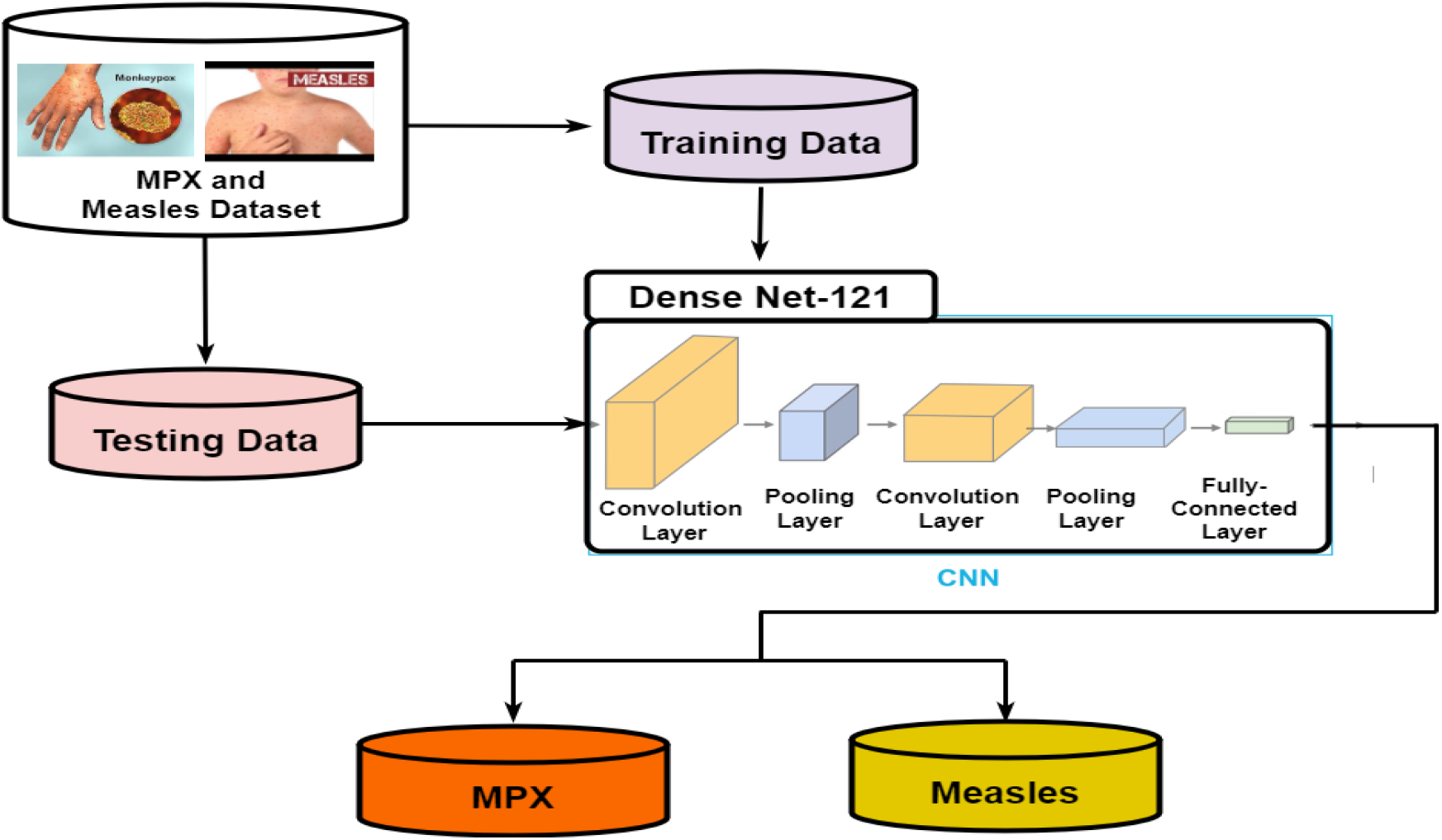
Dense Net-121 model for recognizing MPX.

## 4. Experimental Results and Discussion

To assess the proposed deep learning model, Dense net-121, the confusion matrix, and precision, recall, F1-score, and accuracy metrics were used to evaluate the performance of the proposed model in recognizing MPX within a skin images dataset of MPX and Measles diseases. Moreover, a parallel experiment has been applied using the convolution neural networks (CNN) model on the same dataset to verify the efficiency of the dense net-121model in recognizing MPX images. Table 2 summarizes the initial values of the learning rate and some epochs and the used loss function and optimizer for each deep learning model. Table 3 summarizes the obtained training and testing results of both two models. Figure 3 compares the learning curve and the diminishing loss function curve of the dense net-121 model, and Figure 4 compares the learning curve and the diminishing loss function curve of the CNN model. In addition, Figure 5 compares the confusion matrices of both two models.

**TABLE 2.** Initial settings of applying Dense Net-121 and CNN models.

| Model | learning rate | Loss function | optimizer | No. of epochs | verbose |
| --- | --- | --- | --- | --- | --- |
| DenseNet1-121 | 0.001 | binary_crossentropy | Adam | 15 | 1 |
| CNN | 0.001 | binary_crossentropy | RMS_prop | 50 | 1 |

**TABLE 3.** Training and Testing results of Dense Net-121 and CNN models.

| Training Results |  |  |  |  |  |
| --- | --- | --- | --- | --- | --- |
| Model | Loss value | accuracy | precision | recall | F1_Score |
| DenseNet1-121 | 0.0121 | 1.000 | 1.000 | 1.000 | 0.786 |
| CNN | 0.6568 | 0.648 | 0.648 | 1.00 | 0.786 |
| Testing Results |  |  |  |  |  |
| Model | Loss value | accuracy | precision | recall | F1_Score |
| DenseNet1-121 | 0.354 | 0.937 | 1.00 | 0.8750 | 0.667 |
| CNN | 0.720 | 0.500 | 0.500 | 1.00 | 0.667 |

**Fig. 3.**
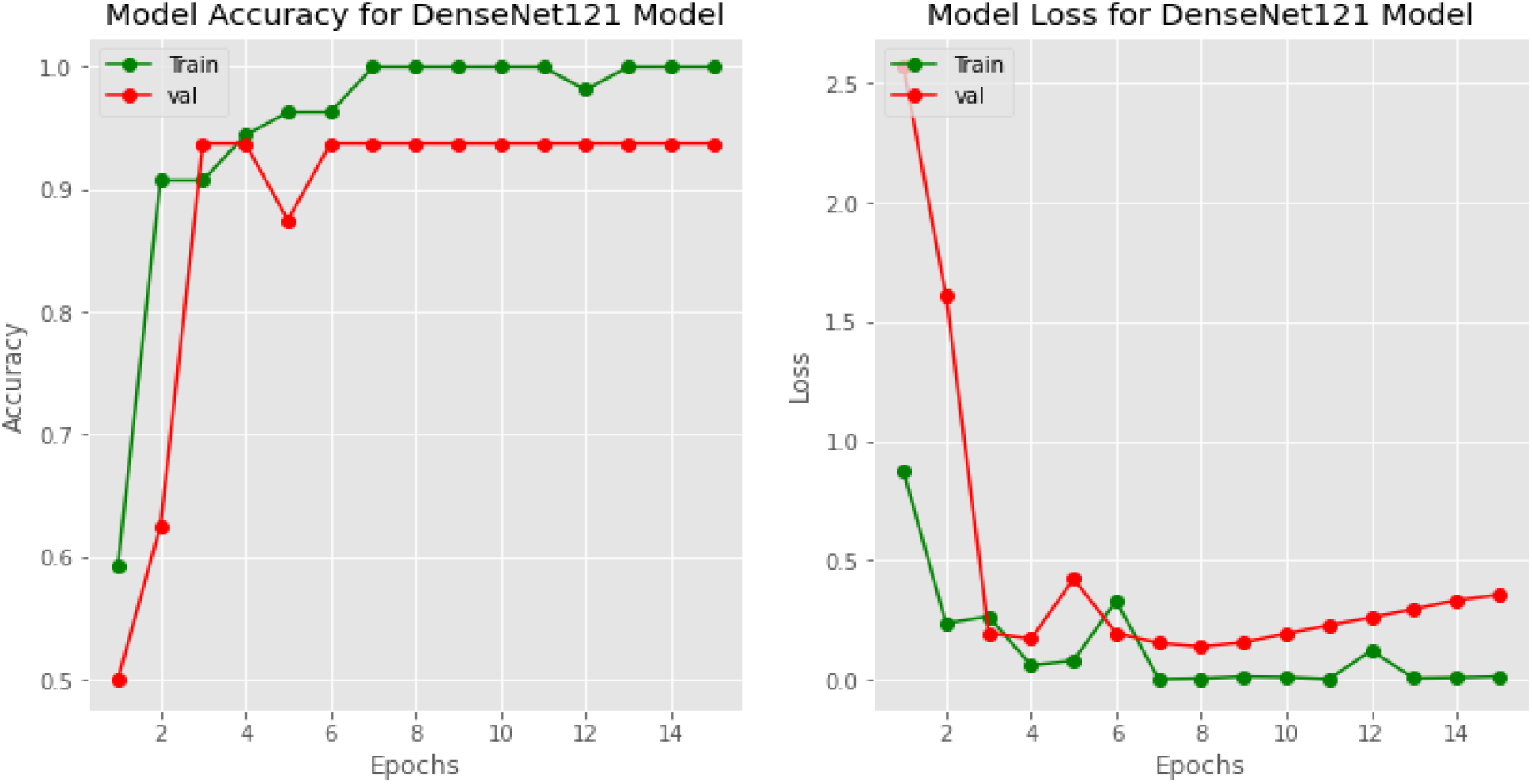
Learning curve and the diminishing loss function curve of the dense net-121 model.

**Fig. 4.**
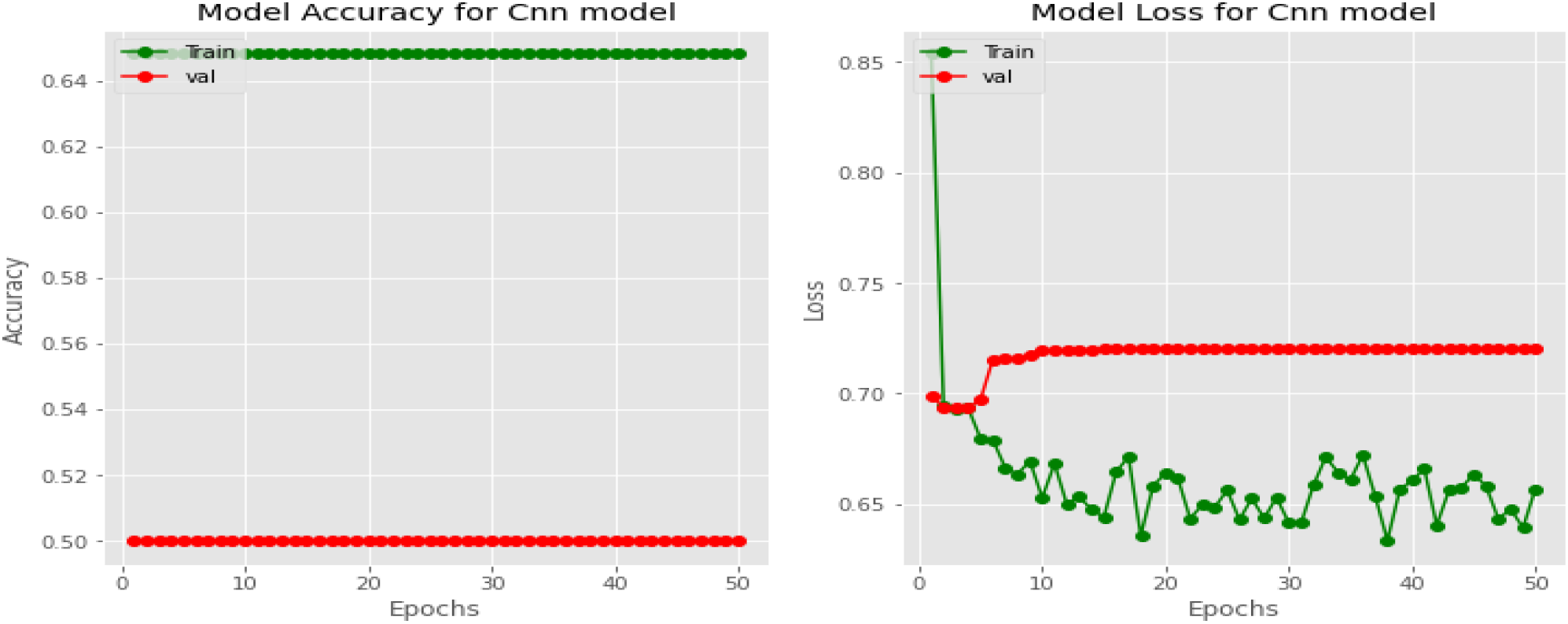
Learning curve and the diminishing loss function curve of CNN model.

**Fig.5.**
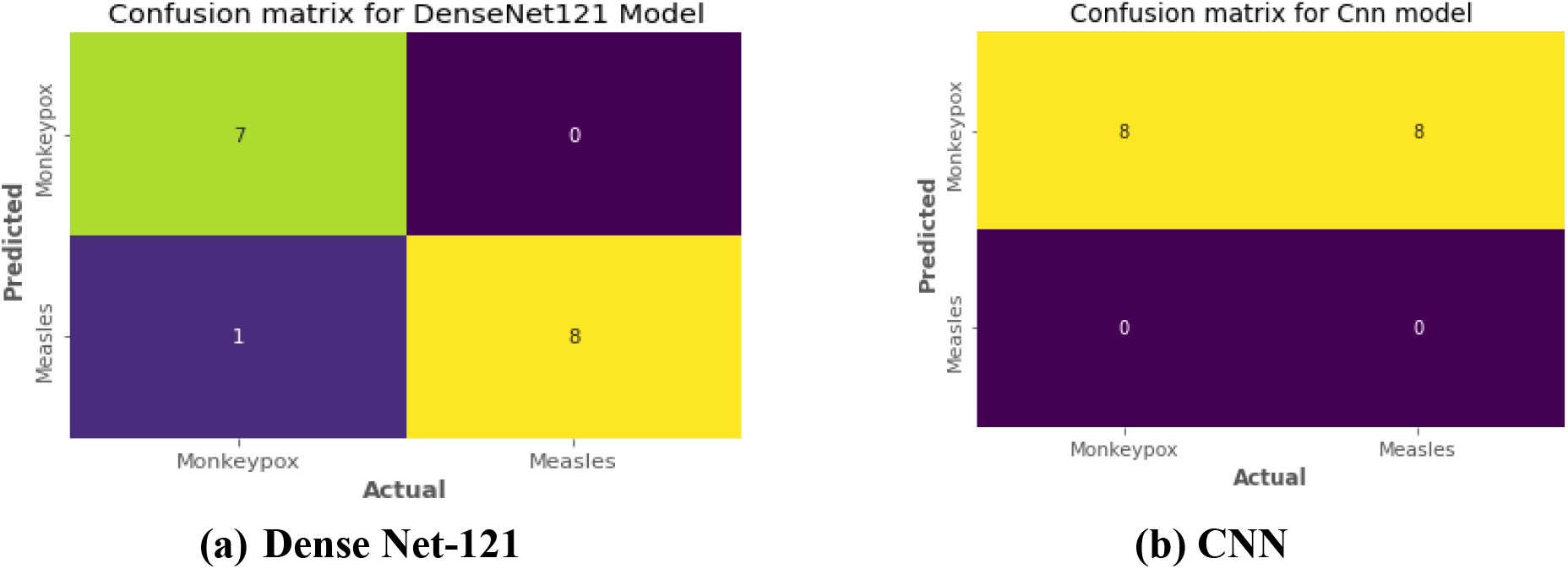
Confusion matrices of (a) Dense Net-121 and (b) CNN models.

The obtained results confirm the superiority of Dense Net-121in recognizing MPX images compared to the classical CNN model regarding training and testing performances, where the Dense Net-121 achieved greater values of accuracy, precision, recall, and F1-score than the CNN model. Moreover, the Dense Net-121 achieved lower loss values in training and testing than CNN as depicted in Figures 3, and 4. In addition, the confusion matrix in Figure 5 (a) clarifies that the dense net-121 succeeded to detect 7-MPX images and failed only to detect one image while performing the testing scenario, this confirms the efficiency of dense net-121 in detecting MPX images compared to CNN model.

## 5. Conclusion

The present study was designed to investigate the efficiency of employing Dens Net-121 as a deep learning model in diagnosing monkeypox through a skin image dataset of MPX and Measles images. The performance of the Dens Net-121 model has been tested and evaluated, then compared with the Convolution Neural Network (CNN) model using a public dataset of skin images of MPX and measles diseases. The most significant finding to emerge from this study is the superiority of the Dense Net-121 model over CNN in diagnosing MPX cases with a testing accuracy of 93%. These findings suggest a role for using deep learning algorithms in diagnosing MPX cases. However, more research trials using other deep learning models are needed to accurately diagnose MPX from big datasets including similar skin and pox diseases such as chicken pox, syphilis, Eczema, etc. This would be a fruitful area for further work.

## Data Availability

All data produced in the present study are available upon reasonable request to the authors

## Notes

### Competing Interest Statement

The authors have declared no competing interest.

### Funding Statement

This study did not receive any funding

